# Are personal health literacy and school health literacy environment important to schoolteachers’ health outcomes?

**DOI:** 10.1101/2024.01.03.24300762

**Authors:** Rongmei Liu, Mingyang Yu, Qiuping Zhao, Junfang Wang, Yuxi Bai, Hui Chen, Xiaomo Yang, Shuaibin Liu, Orkan Okan, Xinghan Chen, Yuhan Xing, Shuaijun Guo

## Abstract

**Background:** While the relationship between an individual’s personal health literacy and health outcomes is well-established, the role of the health literacy environment is often overlooked. This study aimed to examine the associations of personal health literacy and the school health literacy environment with health outcomes among schoolteachers.

**Methods:** A cross-sectional study was conducted in 11 schools in Zhengzhou, Henan, China. Using a self-administered questionnaire, teachers (N=7364) were surveyed collecting data on their sociodemographics, personal health literacy, the school health literacy environment, and four types of health outcomes (health status, health-compromising behaviours, health service use, and healthcare cost). Besides descriptive statistics, a series of logistic regression analyses were conducted.

**Results:** Overall, more than half of teachers (56.9%) had inadequate or problematic health literacy, while more than three-fifths (69.0%) perceived their school health literacy environment was less supportive. Teachers with inadequate health literacy had higher odds of poor health status (odds ratio (OR)=5.79, 95% CI=3.84, 8.73), at least one health-compromising behaviour (OR=2.90, 95% CI=2.29, 3.68), at least one health service use (OR=2.73, 95% CI=2.07, 3.61), and more healthcare cost (OR=2.51, 95% CI=2.00, 3.16) than those with excellent health literacy, after adjusting for sociodemographics and school health literacy environment. Similarly, teachers who perceived low levels of supportive school health literacy environment had higher odds of poor health outcomes (ORs ranging from 1.13 to 1.78), after adjusting for sociodemographics and personal health literacy.

**Conclusion:** Both personal health literacy and school health literacy environment are important to schoolteachers’ health outcomes. Educational programs and organisational change are needed to improve personal health literacy and school environments to improve teachers’ health and well-being.

## INTRODUCTION

Health literacy is a fundamental determinant of health (1). Extant literature has shown that low health literacy is associated with a range of negative health outcomes, including poor health status, health-compromising behaviours, more health service use, and high healthcare costs (2, 3). In the global health promotion agenda of the World Health Organization (WHO) (4), addressing low health literacy is a part of the strategy to tackle health inequities (5). Many national governments and international organisations have integrated health literacy into their health agendas and initiatives and even developed national action plans to improve population health literacy and reduce health disparities (6, 7).

Health literacy is about how an individual manages health information in everyday life in order to make critical health judgments and inform healthy behaviours. Health literacy also includes communicating about health concerns and exchanging health knowledge and health information. However, health literacy goes beyond the individual and should be understood as an interactive outcome between personal health skills and the broad environment where an individual lives and with which it interacts concerning health and wellbeing (8). According to the Australian Commission on Safety and Quality in Health Care (ACSQHC) (9), health literacy has two components: personal health literacy and health literacy environment. Personal health literacy represents one’s ability to access, understand, appraise, and apply health information to maintain and improve personal health, whereas health literacy environment refers to the system’s infrastructure, policies, processes, materials, people, and relationships that influence how people communicate with health information.

Currently, there have been a number of studies that examine the measures, levels, influencing factors, and impact of health literacy across populations (10–13). Findings from a recent European health literacy survey show that low health literacy is prevalent across countries, ranging from 25% in Slovakia to 72% in Germany (14). In China, the 2021 national health literacy survey showed 74.6% of Chinese residents aged 15 to 69 years had low health literacy. However, most existing studies focus on personal health literacy in adults, neglecting the role of health literacy environment, which is an integral part of understanding health literacy in its fullest sense. This is more so true when it comes to children and adolescents and settings relevant to their health and health literacy (15–17). Addressing this issue requires to take into account certain professionals working in school such as teachers.

Teachers play a crucial role in shaping the intellectual and emotional development of school-aged children, but also in the development of children’s health literacy (18). They are uniquely positioned to deliver health education, equipping children with essential health knowledge, attitudes, and behaviours. In addition, they often engage with parents and communities (16), creating a holistic approach to developing children’s health literacy that extends beyond the classroom. Understanding health literacy of teachers is of paramount importance as it not only influences their own health and well-being, but also influences the way they provide health education and promotion programs, thus impacting children’s health literacy (18).

Schools are formal educational organisations and offer infrastructures, resources, and environments that can enable the success of health education and health promotion programs (16), and in particular regarding school health literacy (19, 20). According to the Health Promoting School (HPS) framework (21), there are five key action areas of health promotion in the school setting, which includes: 1) building healthy public policy, 2) creating supportive environments for health, 3) strengthening community action for health, 4) developing personal skills, and 5) re-orienting health services. Empirical evidence shows that all these components have a unique and critical role in equipping both children’s and teacher’s health literacy (22, 23), and fostering their health and well-being (24). Understanding school health literacy environment should be regarded as part of understanding teachers’ health literacy to provide an overall picture of health literacy and its impact on their health outcomes.

Although it has been raised more than two decades ago (18), teachers’ health literacy has only gained increasing attention as part of school health promotion programs in the last decade (18, 25). There have been several national (26, 27) and international studies (28–30) that examine the levels and influencing factors of teachers’ health literacy. Findings from these studies show that teacher’s health literacy is generally low and there are health literacy disparities by sociodemographics such as age, gender, and marital status. However, very few studies have examined the impact of teachers’ health literacy on health outcomes (22). Furthermore, the role of school health literacy environment is often overlooked. To fill these gaps, the present study aimed to investigate the associations of personal health literacy and school health literacy environment with teachers’ health outcomes.

## MATERIALS AND METHODS

### Participants and settings

A cross-sectional study was designed to recruit teachers from 11 schools (five urban and six rural) in Zhengzhou, Henan Province, China, using convenience cluster sampling. Zhengzhou is the capital of Henan, which has a population of 1.28 million in 2022. In brief, two districts (Jingkai District/Zhongmou County) were selected according to their socioeconomic levels, one representing high and the other representing low. Based on previous successful collaborations with schools, we selected five or six schools in each district (Jingkai District: three primary schools and two middle schools; Zhongmou County: four primary schools and two middle schools) according to the appropriateness of survey timing (class time or class break time). At each school, all teaching staff were invited to complete an online self-administered questionnaire via Wenjuanxing. Participants received written online information about the study (e.g., study aim, data collection methods, data storage) in Chinese. Written informed consent was obtained from all respondents prior to filling out the questionnaire. Participants were also informed that they could discontinue their participation at any time. The ethical approval was granted for the study by the Institutional Review Board of Fuwai Central China Cardiovascular Hospital (Ethics ID: 2022-32). Based on previous prevalence studies in Chinese teachers (31) and sample size calculation formula 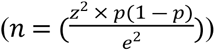 (32), we estimated a sample size of at least 232 in each district (where *p*=0.185, *z*=1.96, *e*=0.05). Considering the potential non-response rate of 30%, the final sample size should be at least 664. Data collection was undertaken between 20 September 2022 and 13 June 2023. We used the Strengthening the Reporting of Observational Studies in Epidemiology (STROBE) checklist (33) as guidelines to ensure the reporting quality of the present study (see S1 Table).

### Questionnaire

A Chinese version questionnaire was designed based on the research purpose to collect information on key sociodemographics, personal health literacy, school health literacy environment, and health outcomes. In total, there were four parts in this questionnaire (Part 1: You and Your Family; Part 2: Your Health Literacy; Part 3: Your School; Part 4: Your Personal Health), with each part having 8 to 12 questions. The average time to complete the survey was 10 minutes.

#### Sociodemographics

We collected socio-demographic information on geographic location (Jingkai District/Zhongmou County), school type (primary/secondary), participants’ sex (male/female), age group (30 years or below/31-39 years/40 years or above), ethnicity (Han/ethnic minorities), marital status (unmarried/married/divorced or widowed), highest educational level (Bachelor or above/Diploma or below), duration of teaching (1-4 years/5-9 years/10-14 years/15-19 years/20-24 years/25 years or more), subject of teaching (literacy/math/English/physics/chemistry/biology/history/geography/politics/ Physical education/music/art/health/other/more than one subject), chronic health conditions (yes/no), health awareness in daily life (very important/not very important), and medical insurance (medical insurance for urban workers/medical insurance for urban and rural residents/rural cooperative medical insurance/commercial medical insurance) (34).

#### Personal health Literacy

The European Health Literacy Population Survey 2019-2021 (HLS_19_ -Q12) was used to measure teachers’ personal health literacy (14). The HLS_19_ -Q12 is a 12-item instrument to assess personal health literacy and was developed by the WHO Action Network on Measuring Population and Organizational Health Literacy (M-POHL) working group. The HLS_19_ -Q12 is based on a matrix with 12 dimensions, which comprises of three health domains (health care, disease prevention, and health promotion) and four aspects of health information management (access, understand, evaluate, and use). Respondents answered each item (e.g., *“On a scale from very easy to very difficult, how easy would you say it is: to find out where to get professional help when you are ill?”*) on a four-point Likert scale (1=very difficult, 4=very easy) concerning the experienced difficulty of each task. The Chinese version the HLS_19_-Q12 has shown excellent reliability and strong validity in the general population of Chinese adults (35). In the present study, our sample had Cronbach’s alpha values of 0.92.

The total score of the HLS_19_ -Q12 was calculated as the percentage (ranging from 0 to 100) of items with valid responses that were answered with “very easy” or “easy” provided that at least 80% of the items contain valid responses (14). If less than 80% of the items contain valid responses, the score was coded as “missing.” Higher scores of the HLS_19_ -Q12 indicate higher levels of health literacy. A categorical variable of the HLS_19_ -Q12 was also calculated based on the recommended cut-off points by WHO M-POHL working group (14) to show the population distribution of health literacy.

#### School health literacy environment

School health literacy environment was assessed by the short form of the organisational health literacy of school questionnaire (OHLS-Q-SF) (20), which was developed based on the school organizational health literacy standards and indicators (36). The OHLS-Q-SF is the short form of the OHLS-Q (37) and consists of eight items that measure health literacy indicators across the whole school environment, including associated processes and structures regarding the promotion of health literacy. Participants answered each item (e.g., “*The design of everyday school life contributes to promoting health literacy at our school”*) on a four-point scale (1 = strongly agree, 2 = agree, 3 = disagree, 4 = strongly disagree). The OHLS-Q-SF showed high reliability (Cronbach’s alpha=0.97) and strong validity (comparative fit index=0.985, Tucker and Lewis’s index of fit=0.978, root mean square error of approximation=0.093 (95% CI=0.088, 0.097) in our sample.

The OHLS-Q-SF total score (ranging 0 to 32) was summed by reversing the code of each item, with higher scores indicating higher levels of organisational health literacy in schools. In keeping with previous studies (38), we used the top 25% as a cut-off to indicate supportive school health literacy environment.

#### Health Outcomes

##### Health status

Health status was assessed using a widely-used general self-report health question (‘*In general, would you say your health is?*’ 1 = poor, 5 = excellent), which has demonstrated strong predictive validity with objective indicators of health and mortality (39). Poor health status was defined if participants answered “poor” or “fair”.

##### Health-compromising behaviours

Health-compromising behaviours were measured by three items derived from previously well-established surveys (40), which included cigarette smoking (“*Are you smoking?*”; 1 = currently; 2 = ever; 3 = never), alcohol drinking (“*Have you had any alcohol in the past 30 days? (more than half a bottle or a can of beer, a small cup of spirit, a glass of wine or yellow wine)*”; 1 = yes; 2 = no), and physical inactivity (“*How many times have you exercised for 30 minutes or more in the past 30 days, such as running, walking, cycling, etc?*”; 1 = almost every day; 2 = several times a week; 3 = several times a month; 4 = almost not at all). Each item was first dichotomised (cigarette smoking: yes = currently or ever/no = never); alcohol drinking: yes/no; physical inactivity: yes = several times a month or almost not at all /no = almost every day or several times a week)) and then a composite measure of health-compromising behaviours was created if participants had at least one health-compromising behaviour.

##### Health service use

Health service use was assessed by four items derived from previously well-established surveys (40), which included emergency service use (“*How many times have you used the emergency service in the last 12 months?*”), general practitioner service use (“*How many times have you been to see a doctor in the last 12 months?*”), hospitalisation (“*How many times have you stayed in a hospital for treatment in the last 12 months?*”), and patient-provider communication (“*How many times have you raised a question during your doctor’s appointment in the last 12 months?*”). Participants answered each item on a four-point scale (1 = 0 times; 2 = 1∼2 times; 3 = 3∼5 times; 4 = 6 or more times). Each item was first dichotomised (yes = 1∼2 times or 3∼5 times or 6 or more times) /no = 0 times) and then a composite measure of health service use was created if participants had used at least one service.

##### Healthcare cost

Healthcare cost was self-reported by participants about the amount of out-of-pocket health expenditure (“*What was your out-of-pocket cost for healthcare (e.g., consultations, medicines, and tests) in the last year?*”) (41). Participants were coded as having high healthcare cost if they spent RMB 1000 or more (41).

### Statistical Analysis

All statistical analyses were performed using Stata 18.0 (StataCorp, Texas, USA). Descriptive statistics were conducted to show the distribution of participants’ characteristics, personal health literacy (continuous and categorical), school health literacy environment (continuous and categorical), and each health outcome. Univariate analysis was used to examine the relationship between participants’ characteristics and levels of personal health literacy and school health literacy environment. Next, a series of logistic regression analyses were conducted to examine the associations between personal health literacy/school health literacy environment and each health outcome. Model 1 was unadjusted. Model 2 was adjusted for all participants’ characteristics (i.e., geographic location, school type, sex, age group, ethnicity, marital status, highest educational level, duration of teaching, subject of teaching, chronic health conditions, health awareness in daily life, and medical insurance). Model 3 was additionally adjusted for school health literacy environment/personal health literacy when examining the association between personal health literacy/school health literacy environment and each health outcome.

#### Missing data

The proportion of respondents with complete data across all study variables was 89.9% (see S1 Appendix 1). To examine the potential impact of missing data, we used multiple imputation by chained equations to reduce the potential bias due to incomplete records (42). The imputation model included all study variables. Based on the percentage of missing data (42), we produced ten imputed datasets and used Rubin’s rules to obtain the final imputed estimates of the parameters of interest. Results using multiply imputed data were reported for all association analyses in the main text.

#### Sensitivity analyses

We also conducted sensitivity analyses to check the robustness of our findings using each indicator of health-compromising behaviours (i.e., cigarette smoking, alcohol drinking, and physical inactivity) and health service use (i.e., emergency service use, general practitioner service use, hospitalisation, and patient-provider communication).

## RESULTS

### Participants’ sociodemographics

In total, 7264 teaching staffs completed the survey, with a response rate of 93.9% (7264/7738). The average age of participants was 35.7±8.3 years (age range: 18 to 68). The majority of participants were recruited from Zhongmou County (82.7%) and primary schools (66.6%). Most participants were female (83.8%), from Han ethnicity (98.4%), and from married families (74.0%). Most teachers had taught more than five years (67.7%). The top three subjects of teaching were literacy (26.2%), math (25.8%) and English (11.1%). The majority (98.7%) of teachers had high health awareness in daily life and had no chronic health conditions (83.3%). The most common type of medical insurance was medical insurance for urban workers (74.8%) (see Table 1).

**Table 1.** Summary of participants’ characteristics. Observed data are shown (n=7264).

| Participants' characteristics | Frequency (%) | Participants' characteristics | Frequency (%) |
| --- | --- | --- | --- |
| Geographic location |  | Subject of teaching |  |
| Jingkai District | 1257 (17.3) | Literacy | 2029 (26.2) |
| Zhongmou County | 6007 (82.7) | Math | 1994 (25.8) |
| School type |  | English | 859 (11.1) |
| Primary school | 4836 (66.6) | Physics | 183 (2.4) |
| Secondary school | 2428 (33.4) | Chemistry | 107 (1.4) |
| Sex |  | Biology | 131 (1.7) |
| Female | 6086 (83.8) | History | 193 (2.5) |
| Male | 1178 (16.2) | Geography | 117 (1.5) |
| Age group |  | Politics | 252 (3.3) |
| 30 years or below | 2448 (33.8) | Physical education | 438 (5.7) |
| 31-39 years | 2609 (36.1) | Music | 264 (3.4) |
| 40 years or above | 2178 (30.1) | Art | 256 (3.3) |
| Ethnicity |  | Health | 38 (0.5) |
| Han | 7150 (98.4) | Other | 226 (2.9) |
| Ethnic minorities | 114 (1.6) | More than one subject | 651 (8.4) |
| Marital status |  | Health awareness in daily life |  |
| Unmarried | 1686 (23.2) | Very important | 7168 (98.7) |
| Married | 5378 (74.0) | Not very important | 96 (1.3) |
| Other* | 200 (2.8) | Chronic health conditions |  |
| Education level |  | No | 6050 (83.3) |
| Bachelor or above | 6338 (87.3) | Yes | 1214 (16.7) |
| Diploma or below | 926 (12.7) | Medical insurance |  |
| Duration of teaching |  | Medical insurance for urban workers | 5365 (74.8) |
| 1-4 years | 2343 (32.3) | Medical insurance for urban and rural residents | 456 (6.4) |
| 5-9 years | 1891 (26.1) | Rural cooperative medical insurance | 1272 (17.7) |
| 10-14 years | 731 (10.1) | Commercial medical insurance | 80 (1.1) |
| 15-19 years | 534 (7.4) |  |  |
| 20-24 years | 816 (11.2) |  |  |
| 25 years or more | 939 (12.9) |  |  |

Note: The category of “other” includes those who are divorced or widowed.

### Distribution of health literacy and health outcomes

Overall, teachers had an average score of 75.17±25.97 in their personal health literacy and scored the school health literacy environment with 25.30±6.35 (Table 2). According to the pre-defined cut-offs in the methods section, we found 13.2% and 30.0% of teachers had excellent and sufficient health literacy respectively. On the other hand, 43.1% and 13.8% of teachers had problematic and inadequate health literacy respectively. Almost one third (31.0%) of teachers perceived their school had a supportive school health literacy environment. Regarding the distribution of health outcomes, we found that 15.0% of teachers had poor health status, 75.2% had at least one health-compromising behaviour, 82.4% had at least one health service use, and 60.7% spent 1000 RMB or more for the out-of-pocket health expenditure.

**Table 2.** Distribution of health literacy and health outcomes. Observed data are shown (n=7264).

| Variable | Frequency (%) / Mean(±SD) |
| --- | --- |
| Personal health literacy (0-100) | 75.17 (25.97) |
| Personal health literacy (categorical) |  |
| Excellent | 901 (13.2) |
| Sufficient | 2046 (30.0) |
| Problematic | 2942 (43.1) |
| Inadequate | 942 (13.8) |
| School health literacy environment (8-32) | 25.30 (6.35) |
| School health literacy environment (categorical) |  |
| Supportive | 2217 (31.0) |
| Not supportive | 4931 (69.0) |
| Health status |  |
| Good | 6178 (85.0) |
| Poor | 1086 (15.0) |
| Cigarette smoking |  |
| Never | 6959 (95.8) |
| Ever or current | 305 (4.2) |
| Alcohol drinking |  |
| No | 5751 (79.2) |
| Yes | 1513 (20.8) |
| Physical inactivity |  |
| No | 2481 (34.2) |
| Yes | 4783 (65.8) |
| At least one health-compromising behaviour |  |
| No | 1799 (24.8) |
| Yes | 5465 (75.2) |
| Emergency service |  |
| No | 5862 (81.8) |
| Yes | 1302 (18.2) |
| General practitioner service |  |
| No | 1548 (21.5) |
| Yes | 5641 (78.5) |
| Hospitalisation |  |
| No | 5387 (74.9) |
| Yes | 1807 (25.1) |
| Patient-provider communication |  |
| No | 1799 (25.1) |
| Yes | 5370 (74.9) |
| At least one health service use |  |
| No | 1271 (17.6) |
| Yes | 5949 (82.4) |
| Healthcare cost |  |
| <1000 RMB | 2847 (39.3) |
| ≥1000 RMB | 4396 (60.7) |

SD, standard deviation.

### Distribution of health literacy by participants’ characteristics

Table 3 shows the relationship between personal health literacy/school health literacy environment and participants’ characteristics. Teachers had high personal health literacy scores if they were from primary schools, female, younger, unmarried, taught physical education, had high health awareness in daily life, no chronic health conditions, and medical insurance for urban and rural residents. Similarly, teachers perceived high levels of school health literacy environment if they were from primary schools, younger, from Han ethnicity backgrounds, unmarried, taught physical education, had high health awareness in daily life, no chronic health conditions, and medical insurance for urban and rural residents.

**Table 3.** Distribution of health literacy by participants’ characteristics, using imputed samples (n=7264).

| Participants' characteristics | Mean(±SD) | |
| --- | --- | --- |
|  | Personal health literacy | School health literacy environment |
| Geographic location |  |  |
| Jingkai District | 73.47 (72.00, 74.93) <sup>a</sup> | 25.50 (25.15, 25.85) <sup>a</sup> |
| Zhongmou County | 74.67 (73.98, 75.35) <sup>a</sup> | 25.21 (25.05, 25.38) <sup>a</sup> |
| School type |  |  |
| Primary school | 75.49 (74.75, 76.22) <sup>a</sup> | 25.73 (25.56, 25.90) <sup>a</sup> |
| Secondary school | 72.42 (71.30, 73.54) <sup>b</sup> | 24.33 (24.06, 24.60) <sup>b</sup> |
| Sex |  |  |
| Female | 74.61 (73.96, 75.25) <sup>a</sup> | 25.29 (25.13, 25.45) <sup>a</sup> |
| Male | 73.70 (71.96, 75.44) <sup>a</sup> | 25.14 (24.77, 25.52) <sup>a</sup> |
| Age group |  |  |
| 30 years or below | 80.48 (79.55, 81.42) <sup>a</sup> | 26.34 (26.10, 26.57) <sup>a</sup> |
| 31-39 years | 74.48 (73.47, 75.49) <sup>b</sup> | 25.40 (25.15, 25.64) <sup>b</sup> |
| 40 years or above | 67.69 (66.45, 68.92) <sup>c</sup> | 23.90 (23.62, 24.19) <sup>c</sup> |
| Ethnicity |  |  |
| Han | 74.51 (73.89, 75.13) <sup>a</sup> | 25.28 (25.14, 25.43) <sup>a</sup> |
| Ethnic minorities | 71.24 (65.84, 76.64) <sup>a</sup> | 23.96 (22.56, 25.35) <sup>b</sup> |
| Marital status |  |  |
| Unmarried | 79.23 (78.04, 80.42) <sup>a</sup> | 25.97 (25.69, 26.26) <sup>a</sup> |
| Married | 73.11 (72.38, 73.83) <sup>b</sup> | 25.05 (24.87, 25.22) <sup>b</sup> |
| Other* | 70.66 (66.67, 74.66) <sup>b</sup> | 25.08 (24.12, 26.04) <sup>ab</sup> |
| Education level |  |  |
| Bachelor or above | 74.43 (73.77, 75.09) <sup>a</sup> | 25.20 (25.04, 25.36) <sup>a</sup> |
| Diploma or below | 74.67 (72.86, 76.49) <sup>a</sup> | 25.70 (25.31, 26.09) <sup>a</sup> |
| Duration of teaching |  |  |
| 1-4 years | 79.98 (79.01, 80.94) <sup>a</sup> | 26.57 (26.34, 26.79) <sup>a</sup> |
| 5-9 years | 76.13 (74.93, 77.33) <sup>b</sup> | 25.37 (25.07, 25.66) <sup>b</sup> |
| 10-14 years | 73.29 (71.31, 75.26) <sup>c</sup> | 24.80 (24.32, 25.29) <sup>c</sup> |
| 15-19 years | 71.57 (69.12, 74.02) <sup>c</sup> | 24.23 (23.65, 24.81) <sup>cd</sup> |
| 20-24 years | 68.58 (66.61, 70.54) <sup>d</sup> | 24.36 (23.91, 24.82) <sup>cd</sup> |
| 25 years or more | 65.06 (63.19, 66.93) <sup>e</sup> | 23.54 (23.10, 23.98) <sup>e</sup> |
| Subject of teaching |  |  |
|  | Personal health literacy | School health literacy environment |
| Literacy | 75.02 (73.84, 76.21)<sup>ac</sup> | 25.28 (25.00, 25.56)<sup>ace</sup> |
| Math | 73.36 (72.16, 74.56)<sup>ad</sup> | 25.39 (25.12, 25.66)<sup>c</sup> |
| English | 73.92 (72.07, 75.76)<sup>ad</sup> | 24.82 (24.35, 25.29)<sup>ab</sup> |
| Physics | 66.50 (62.12, 70.88)<sup>b</sup> | 23.81 (22.82, 24.80)<sup>b<sup>g</sup>f</sup> |
| Chemistry | 68.39 (62.81, 73.98)<sup>b<sup>d</sup>f</sup> | 23.00 (21.58, 24.43)<sup>g</sup> |
| Biology | 76.13 (71.46, 80.80)<sup>ag</sup> | 26.04 (25.15, 26.93)<sup>ceh</sup> |
| History | 72.35 (68.12, 76.59)<sup>af</sup> | 24.02 (23.03, 25.01)<sup>bg</sup> |
| Geography | 74.80 (70.23, 79.38)<sup>afi</sup> | 24.33 (23.07, 25.59)<sup>acfg</sup> |
| Politics | 73.18 (69.53, 76.83)<sup>afj</sup> | 25.03 (24.16, 25.90)<sup>acf</sup> |
| Physical education | 82.47 (79.98, 84.96)<sup>ek</sup> | 27.05 (26.50, 27.61)<sup>dh</sup> |
| Music | 79.24 (76.00, 82.49)<sup>gikl</sup> | 26.53 (25.68, 27.39)<sup>dhi</sup> |
| Art | 76.41 (72.89, 79.93)<sup>hijm</sup> | 25.83 (24.98, 26.67)<sup>cei</sup> |
| Health | 72.38 (61.05, 83.70)<sup>ablm</sup> | 24.56 (21.87, 27.25)<sup>acfgi</sup> |
| Other | 74.05 (70.02, 78.08)<sup>af<sup>lm</sup></sup> | 25.51 (24.49, 26.53)<sup>aci</sup> |
| More than one subject | 73.40 (71.25, 75.55)<sup>afm</sup> | 24.80 (24.27, 25.32)<sup>acf</sup> |
| Health awareness in daily life |  |  |
| Very important | 74.67 (74.05, 75.28)<sup>a</sup> | 25.30 (25.16, 25.45)<sup>a</sup> |
| Not very important | 59.03 (52.30, 65.76)<sup>b</sup> | 22.23 (20.74, 23.71)<sup>b</sup> |
| Chronic health conditions |  |  |
| No | 76.96 (76.33, 77.58)<sup>a</sup> | 25.79 (25.64, 25.95)<sup>a</sup> |
| Yes | 62.02 (60.25, 63.78)<sup>b</sup> | 22.63 (22.24, 23.03)<sup>b</sup> |
| Medical insurance |  |  |
| Medical insurance for urban workers | 73.77 (73.06, 74.48)<sup>a</sup> | 24.98 (24.80, 25.15)<sup>a</sup> |
| Medical insurance for urban and rural residents | 79.28 (76.79, 81.77)<sup>b</sup> | 26.55 (26.03, 27.07)<sup>b</sup> |
| Rural cooperative medical insurance | 75.49 (73.99, 76.99)<sup>c</sup> | 26.01 (25.68, 26.34)<sup>bc</sup> |
| Commercial medical insurance | 76.64 (70.87, 82.42)<sup>abc</sup> | 25.15 (23.62, 26.68)<sup>abc</sup> |

SD, standard deviation. Note: The category of “other” includes those who are divorced or widowed.
Distribution of health literacy with the same character indicates no statistical difference between groups.

### Associations between health literacy with health outcomes

Compared with those who had excellent health literacy, teachers with inadequate health literacy had higher odds of poor health status (odds ratio (OR)=5.79, 95% CI=3.84, 8.73), at least one health-compromising behaviour (OR=2.90, 95% CI=2.29, 3.68), at least one health service use (OR=2.73, 95% CI=2.07, 3.61), and more healthcare cost (OR=2.51, 95% CI=2.00, 3.16), after adjusting for all covariates and school health literacy environment (see Table 4). Similarly, teachers who perceived their school to have low levels of supportive school health literacy environment had higher odds of poor health status (OR=1.62, 95% CI=1.32, 1.99), at least one health-compromising behaviour (OR=1.39, 95% CI=1.22, 1.58), at least one health service use (OR=1.78, 95% CI=1.54, 2.06), and more healthcare cost (OR=1.13, 95% CI=1.00, 1.27), after adjusting for all covariates and personal health literacy.

**Table 4.** Associations between health literacy and health outcomes, using imputed samples (n=7264).

|  | Model 1 | Model 2 | Model 3 |
| --- | --- | --- | --- |
| *Association with poor health status* |  |  |  |
| Personal health literacy |  |  |  |
| Excellent | Ref | Ref | Ref |
| Sufficient | 1.94 (1.30, 2.91) | 1.69 (1.12, 2.54) | 1.39 (0.91, 2.11) |
| Problematic | 5.30 (3.67, 7.64) | 3.93 (2.71, 5.69) | 3.07 (2.08, 4.51) |
| Inadequate | 12.36 (8.46, 18.07) | 7.73 (5.21, 11.46) | 5.79 (3.84, 8.73) |
| School health literacy environment |  |  |  |
| Supportive | Ref | Ref | Ref |
| Not supportive | 3.11 (2.59, 3.73) | 2.36 (1.95, 2.86) | 1.62 (1.32, 1.99) |
| *Association with at least one health-compromising behaviour* |  |  |  |
| Personal health literacy |  |  |  |
| Excellent | Ref | Ref | Ref |
| Sufficient | 1.87 (1.59, 2.20) | 1.93 (1.63, 2.28) | 1.71 (1.43, 2.04) |
| Problematic | 2.50 (2.13, 2.93) | 2.81 (2.38, 3.32) | 2.38 (1.99, 2.85) |
| Inadequate | 3.08 (2.50, 3.79) | 3.55 (2.84, 4.43) | 2.90 (2.29, 3.68) |
| School health literacy environment |  |  |  |
| Supportive | Ref | Ref | Ref |
| Not supportive | 1.71 (1.53, 1.91) | 1.80 (1.61, 2.03) | 1.39 (1.22, 1.58) |
| *Association with at least one health service use* |  |  |  |
| Personal health literacy |  |  |  |
| Excellent | Ref | Ref | Ref |
| Sufficient | 2.25 (1.88, 2.68) | 2.12 (1.77, 2.54) | 1.72 (1.42, 2.08) |
| Problematic | 3.58 (3.01, 4.25) | 3.07 (2.57, 3.67) | 2.30 (1.90, 2.80) |
| Inadequate | 5.00 (3.90, 6.42) | 3.93 (3.02, 5.10) | 2.73 (2.07, 3.61) |
| School health literacy environment |  |  |  |
| Supportive | Ref | Ref | Ref |
| Not supportive | 2.55 (2.25, 2.89) | 2.31 (2.02, 2.63) | 1.78 (1.54, 2.06) |
| **Association with healthcare cost more than 1000 RMB** |  |  |  |
| Personal health literacy |  |  |  |
| Excellent | Ref | Ref | Ref |
| Sufficient | 1.41 (1.20, 1.65) | 1.31 (1.11, 1.55) | 1.25 (1.05, 1.49) |
| Problematic | 2.04 (1.75, 2.37) | 1.64 (1.40, 1.93) | 1.55 (1.30, 1.84) |
| Inadequate | 3.67 (3.00, 4.49) | 2.71 (2.18, 3.36) | 2.51 (2.00, 3.16) |
| School health literacy environment |  |  |  |
| Supportive | Ref | Ref | Ref |
| Not supportive | 1.58 (1.43, 1.75) | 1.35 (1.21, 1.50) | 1.13 (1.00, 1.27) |

Model 1: Unadjusted; Model 2: Adjusted for geographic location, school type, sex, age group, ethnicity, marital status, education level, duration of teaching, subject of teaching, health awareness in daily life, chronic health conditions, and medical insurance; Model 3: Additionally adjusted for school health literacy environment when examining the impact of personal health literacy or additionally adjusted for personal health literacy when examining the impact of school health literacy environment.

We also found similar results when examining the associations of personal health literacy and school health literacy environment with each indicator of health-compromising behaviours (see S2 Appendix 2) and health service use (see S3 Appendix 3).

## DISCUSSION

### Summary of key findings in the present study

Using a cross-sectional study design, we examined the relationships between personal health literacy, school health literacy environment, and a range of health outcomes among Chinese schoolteachers. This was also the first study in Asia to assess organizational health literacy in school settings. Specifically, there were three main findings in the present study: (1) there was a high proportion (56.9%) of schoolteachers with inadequate or problematic health literacy; (2) the study found sociodemographic differences (e.g., age, marital status, teaching school type) in teacher’s personal health literacy and within the school health literacy environment; (3) Both personal health literacy and school health literacy environment were associated with health status, health-compromising behaviours, health service use, and healthcare cost.

Consistent with findings from previous research (28, 30), we found that low health literacy was prevalent (56.9%) in our sample when using the HLS_19_ -Q12. As shown in the 2012 Chinese national health literacy survey (31), 81.5% of Chinese teachers had low health literacy. Internationally, in a study conducted in Turkey, Yilmazel and Cetinkaya (28) used the 6-item Newest Vital Sign (NVS) to measure 500 primary and secondary teachers’ health literacy and found 73.8% of teachers had low health literacy. Denuwara and Gunawardena (30) found that 32.5% of secondary teachers had low health literacy in Sri Lanka when using the 47-item European Health Literacy Survey Questionnaire (HLS-EU-Q47), whereas Rahimi and Tavassoli found 48.3% to 60% of primary teachers had low health literacy in Iran using the 36-item Test of Functional Health Literacy in Adults (TOFHLA). While these studies use different instruments to measure health literacy, there is consistent evidence showing the pressing need to improve schoolteachers’ health literacy.

Measuring health literacy and its influencing factors are important to provide insights for and inform intervention development. We found that teachers’ health literacy levels varied by school type, age group, marital status, duration of teaching, subject of teaching, health awareness in daily life, chronic health condition status, and medical insurance type. Most of these findings align with previous similar studies (28, 30), showing that teachers tended to have low levels of health literacy if they were older, had longer duration of teaching, and chronic health conditions. Future intervention studies may consider these characteristics when targeting schoolteachers’ health literacy. However, we did not find differences of health literacy by geographic location, sex, and highest educational level, which was contrary to previous research (28, 29). One possible reason could be the convenience sampling approach, which we recruited teachers from two districts in one city, thus contributing to homogeneity of our samples (i.e., 82.7% was from Zhongmou County, 83.8% were female, and 87.3% had Bachelor’s degree or above). Further research is needed to use more representative samples to investigate these influencing factors of teachers’ health literacy, given their potential roles as social determinants of health.

Compared to previous studies that examined teachers’ health literacy (28–30, 43), we added evidence on the impact of health literacy on teachers’ health outcomes. We found teachers with lower health literacy were likely to have poorer health status, more health-compromising behaviours, more health service use, and more healthcare costs. These findings are consistent with previous systematic reviews (3, 44) and population-based studies (41, 45). The potential pathways linking health literacy with health outcomes include both personal factors (e.g., self-efficacy, health knowledge, health awareness) and system factors (e.g., complexity of healthcare systems, social support, provider competence). Our findings suggest promoting teachers’ health literacy skills may have the potential to improve their health outcomes. As shown in the recent evidence base (46, 47), health literacy interventions can lead to improved health outcomes in the general population. However, evidence is scarce regarding health literacy interventions targeting teachers. Future experimental research is needed to support the protective role of health literacy in predicting teachers’ health and wellbeing, and what role teachers’ health literacy plays in context of enhancing school-aged children’s health literacy and health outcomes.

The present study also extends current literature by examining the levels, influencing factors, and impacts of the school health literacy environment. In keeping with the distribution of personal health literacy among teachers, we found similar patterning of school health literacy environment. Teachers perceived lower levels of supportive school health literacy environment if they were older, from ethnic minority backgrounds, married, had longer duration of teaching, taught subjects other than physical education and biology, had low health awareness, had chronic health conditions, and had medical insurance for urban workers. We also found lower levels of supportive school health literacy environment were associated with poor health status, more health-compromising behaviours, more health service use, and more healthcare costs among teachers. Consistent with previous research (22, 23), our findings show that school health literacy environment is an important situational factor to influence teachers’ health and well-being, with potential direct and indirect pathways through personal health literacy (48), social support (22), and mental health (49). Also, echoing the HPS framework and other similar concepts (e.g., HeLit-Schools in Germany (20), HealthLit4Kids in Australia (50)), our findings suggest that school health literacy environment should be included as an essential part of heath literacy interventions to improve teachers’ health and well-being.

## Limitations

There were several limitations that should be noted. First, we used the convenience sampling to recruit schoolteachers from two districts of Zhengzhou, Henan Province. Our findings may not generalise to other geographic regions or populations. There is a need for future research to recruit more representative samples to replicate our findings. Second, measurement errors may exist for self-report instruments. In the present study, we used previously validated items or instruments where possible to enhance the validity and reliability of our measures, thus minimising the self-report bias. Third, our findings are based on the cross-sectional study design, therefore we could not establish causality. Further research using longitudinal or experimental designs is needed to confirm our findings. Finally, we did not consider other social environments (e.g., mass media, healthcare, family) that may contribute to teachers’ health outcomes, given our focus was on schools where they work. Given that health literacy is a context-specific and relational concept, future research may explore other health literacy environments and their impacts on health outcomes.

### Implications for future research and practice

Findings from the present study shed light on the critical intersections between personal health literacy, the school health literacy environment, and various health outcomes among Chinese schoolteachers. Future research may consider using similar approaches to examining the role of personal health literacy and health literacy environment in other populations (e.g., children, elderly, patients) or settings (e.g., workplace, hospitals, markets). Also, it would be valuable to explore these relationships from a more precise perspective. For example, the HLS_19_ -Q12 used to measure personal health literacy has three health domains (health care, disease prevention, and health promotion) and four aspects of health information management (access, understand, evaluate, and use). Variations may exist between these sub-components for the relationships with health outcomes.

Given that more than half of teachers have inadequate or problematic health literacy, it is imperative for governments and schools to design and implement interventions to improve teachers’ health literacy. Future research could explore the feasibility and effectiveness of incorporating health literacy education into pre-service and in-service teacher training to equip educators with sufficient health knowledge and skills (51). The identified sociodemographic differences in personal health literacy and school health literacy environment also can provide insights into specific demographic groups to whom the intervention should target. To improve health outcomes for schoolteachers, both educational programs and organisational change are needed to improve personal health literacy and school environments. There is increasing attention to programs that use a whole-of-school approach to promoting teachers’ health literacy and improving their health outcomes. For example, the HeLit-Schols project (20) and the HealthLit4Kids project (50) both highlight the need of organisational change to sustainably promote health literacy for children and all staff in school settings.

## CONCLUSIONS

This study examined the relationship between health literacy and teachers’ health outcomes from two aspects: personal health literacy and school health literacy environment. We found that low health literacy was common in Chinese schoolteachers. The sociodemographic differences observed in both personal health literacy and school health literacy environment highlight the importance of tailoring interventions to address specific needs for subgroups of the diverse teacher population. Our findings also show that both personal health literacy and school health literacy environment are important to teachers’ health status, health behaviours, health service use and healthcare cost. Promoting health literacy at both individual and organizational levels has the potential to improve population health and reduce health disparities.

## Data Availability

Data cannot be shared publicly because of risk of violating privacy. Data are available from the Institutional Review Board of Fuwai Central China Cardiovascular Hospital (contact via Mr Taibing Fan, Phone: +86-0371-58680341) for researchers who meet the criteria for access to confidential data.

## Supporting information

S1 Table. STROBE checklist.

S1 Appendix 1. Missing data.

S2 Appendix 2. Associations between health literacy and each indicator of health behaviours.

S3 Appendix 3. Associations between health literacy and each indicator of health service use.

## Aknowledgments

We thank all the schools and teachers who participated in the study.

## Authors’ contributions

QZ, RL, MY and SG contributed to the study design, drafted the initial manuscript, critically reviewed the manuscript for important intellectual content. RL and MY was responsible for the field work, data collection and quality control. SG performed statistical analysis and interpreted results. JW, YB, HC, XY, SL, XC, YX and OO contributed to the study design and critically reviewed the manuscript for important intellectual content. All authors read and approved the final manuscript.

## Funding

This research is supported by c (SB201903022) and Henan Province Science and Technology Research Program Soft Science Project (RKX202201002).

## Availability of data and materials

The datasets used during the current study are available from the corresponding author on reasonable request.

## Completing interests

The authors declare that they have no competing interests.

## Notes

### Competing Interest Statement

The authors have declared no competing interest.

### Author Declarations

Participants received written online information about the study (e.g., study aim, data collection methods, data storage) in Chinese. Written informed consent was obtained from all respondents prior to filling out the questionnaire. Participants were also informed that they could discontinue their participation at any time. The ethical approval was granted for the study by the Institutional Review Board of Fuwai Central China Cardiovascular Hospital (Ethics ID: 2022-32).

